# Diurnal dynamics and psychobiological regulation of cell-free mitochondrial and nuclear DNA in human saliva

**DOI:** 10.1101/2025.10.13.25337887

**Authors:** Alexander Behnke, David Shire, Samantha Leonard, Aidan Park, Lauren Petri, Anjali Goyal, Clemens Kirschbaum, Martin Picard, Caroline Trumpff

## Abstract

Cell-free mitochondrial DNA (cf-mtDNA) has emerged as a dynamic molecular signal responsive to psychological stress and a potential biomarker of stress-related physiological adaptations. While previous work has established the acute stress reactivity of cf-mtDNA in saliva, little is known about its diurnal regulation. In this intensive-sampling study, we characterized the diurnal dynamics of saliva cf-mtDNA across one weekday and one weekend day in healthy adults (*N* = 25, 52% female, 826 samples). Saliva was collected at awakening, during the first hour post-awakening, and at hourly intervals throughout the day. Using a quantitative PCR-based assay, we observed a robust cf-mtDNA awakening response, with concentrations peaking approximately 45 minutes after waking (2.5-fold change, Cohen’s *d* = 0.90), followed by a second peak at 180 minutes post-awakening (5.7-fold change, *d* = 1.70) and relatively stable levels thereafter. Saliva cf-mtDNA closely tracked cell-free nuclear DNA (cf-nDNA) across time points (*r*_S_ = .85), suggesting shared release mechanisms. Diurnal cf-mtDNA showed limited correspondence with cortisol and other hormones in saliva. Psychosocial stress indicators—including daily hassles, lack of social support, negative emotional affect, trait anxiety, fatigue, and depressive symptoms—were associated with higher awakening cf-mtDNA levels, a diminished awakening response, and lower diurnal variability. These findings suggest that saliva cf-mtDNA exhibits a diurnal rhythm and is sensitive to psychosocial stress exposure. By establishing its diurnal patterns and individual-level variability, this study advances saliva cf-mtDNA as a promising non-invasive biomarker to dynamically capture stress-related mitochondrial signaling.

## Introduction

Chronic psychosocial stress is a key risk factor for a range of adverse health outcomes, including accelerated aging, impaired immune function, and increased susceptibility to mental and physical disease [1, 2]. Although acute stress responses are adaptive, chronic and repeated stress exposure can alter physiological processes in complex, yet not fully understood ways [1, 3, 4]. One promising molecular signal implicated in stress adaptation processes and stress pathophysiology is *cell-free mitochondrial DNA* (cf-mtDNA)—fragments or entire copies of the mitochondrial genome present in the circulation and extracellular space, either freely or encapsulated in vesicles and organelle-like structures [5–10].

Cf-mtDNA levels in blood rise within minutes after exposure to acute physical and psychosocial stressors, suggesting that it may act as a rapidly inducible biomarker of psychophysiological stress responses [11–13, 10, 14]. Furthermore, blood cf-mtDNA levels are observed to fluctuate several-fold in the timescale of weeks, which has been linked in parts to stress exposure and related negative consequences, such as depressive symptoms [11, 12, 15–19].

These dynamics mirror the release patterns of established stress mediators, such as the steroid hormone cortisol, giving rise to the hypothesis that cf-mtDNA might serve as a valuable biomarker for monitoring acute and chronic stress, offering insights into stress-related adaptations at the nexus of energy metabolism and immune processes [6, 9, 20–23]. Notably, cf-mtDNA can be accessed non-invasively in human saliva, which has sparked growing interest in saliva cf-mtDNA dynamics as an easy-to-access stress biomarker [6, 24]. Recent findings indicate that saliva cf-mtDNA responds even more sensitively to a psychosocial stress paradigm than blood cf-mtDNA, emphasizing its potential utility in stress research [13, 10].

Additionally, studies also revealed a rhythmic pattern of blood cf-mtDNA release, similar to other stress biomarkers like cortisol, but with distinct rhythmic characteristics [9, 11]. Importantly, in an initial study in two men performing four daily saliva collections over 53–60 consecutive days, we observed that saliva cf-mtDNA undergoes a pattern of diurnal rhythmicity, characterized by an awakening response, with marked 2- to 3-fold average increases in the first 30-45 minutes after waking (with large intra-individual, day-to-day differences), followed by rather stable levels throughout the day [25]. These preliminary findings raise the possibility that saliva cf-mtDNA dynamics follow daily rhythms, conceivably influenced by both stress exposure [25] and other biological and lifestyle factors, such as age, sex, BMI, sleep patterns, and physical activity [9, 19, 26].

The present study aimed to investigate saliva cf-mtDNA as a potential stress biomarker through characterizing its diurnal dynamics in healthy individuals across multiple time points throughout two different days. We examined the cf-mtDNA awakening response and its associations with psychosocial stress, individual characteristics, and lifestyle factors. In addition, we compared the awakening responses of cf-mtDNA and cortisol in saliva (Figure 1A), as cell-based experiments have shown that cortisol stimulation triggers mtDNA release from human fibroblasts [12], and our previous study suggested coordinated diurnal dynamics of the two markers [25]. Elucidating the psychobiological regulation of diurnal cf-mtDNA dynamics will advance the methodological consolidation of saliva cf-mtDNA assessments in stress research, and it will shed light on potential physiological adaptations to stress and related negative health conditions.

**Figure 1.**
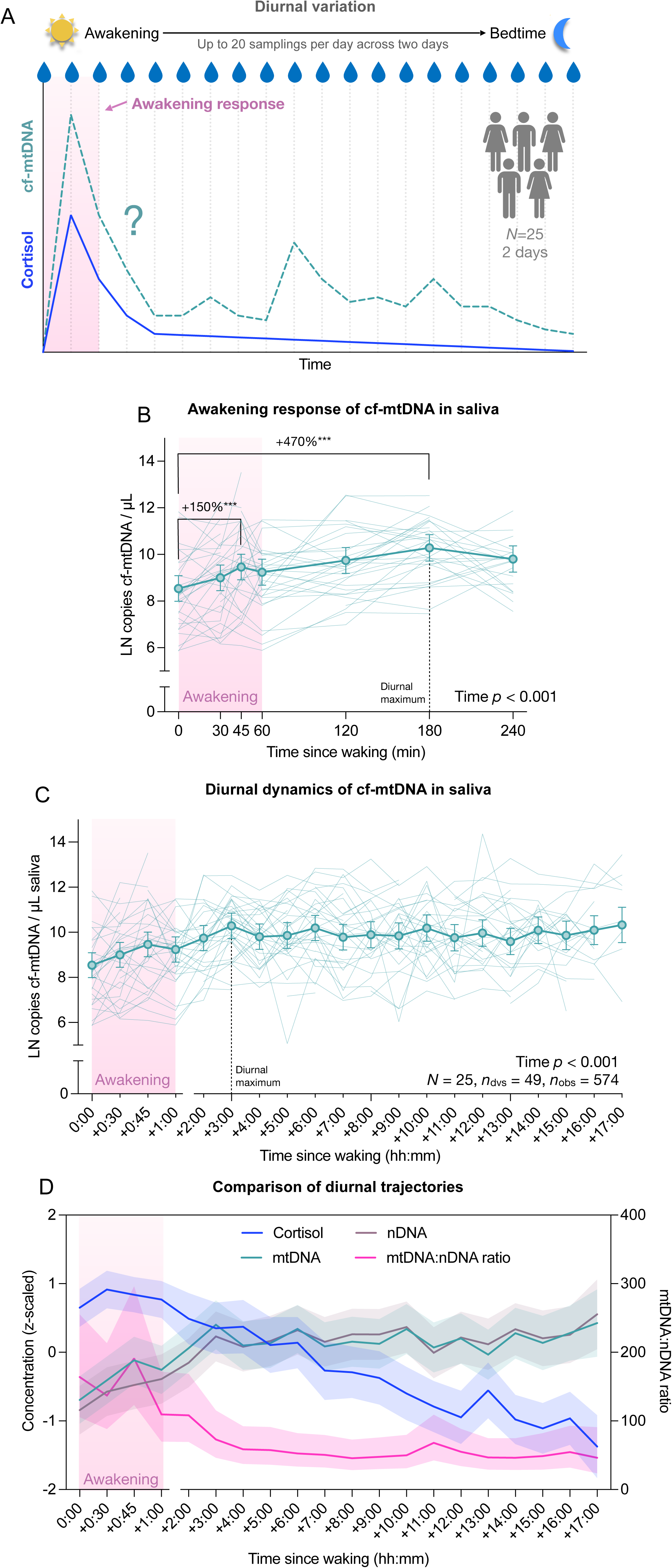
Saliva cf-mtDNA diurnal variation and awakening response. **(A)** Overview of the experimental design. Healthy participants (*N* = 25) collected up to 20 saliva samples over the course of one weekday and one weekend day for saliva cell-free DNA and steroid hormones (including cortisol) measurements. **(B)** Diurnal saliva cell-free mitochondrial DNA (cf-mtDNA) trajectories for the awakening response, and **(C)** for all time points. **(D)** Diurnal trajectories of cortisol (blue), cell-free mitochondrial DNA (cf-mtDNA, green), and cell-free nuclear DNA (cf-nDNA, violet), as well as the mtDNA-to-nDNA ratio (pink) are compared. Values are presented as (B,C) marginal means from mixed-effects models with 95% confidence intervals (bold opaque lines and symbols) and individual trajectories (thin transparent lines), (D, left y-axis) *z*-scaled natural log-transformed marginal mean concentrations of cortisol, mtDNA, and nDNA with 95% confidence intervals, and (D, right y-axis) mtDNA-to-nDNA ratio, with 95% confidence interval. Effect sizes and *p*-values from (B,C) linear mixed-effects models and selected Tukey’s post hoc tests, *** *p*_Tukey_ < 0.001, two-tailed. For clarity, only the post hoc comparisons showing the greatest changes have been plotted.

## Materials and Methods

### Study cohort

A total of 25 healthy adults (52% female; age: *M* = 31.1, *SD* = 8.0 years; BMI: *M* = 23.9, *SD* = 3.1 kg/m²; 96% non-smokers) participated in this study (see Table 1 for participant characteristics). Participants were recruited from the local Columbia University Irving Medical Center (CUIMC) community, using a stratification scheme to balance the cohort’s diversity in terms of ethnicity and regional ancestry. Eligibility was determined based on inclusion/exclusion criteria (see Supplementary Material) and participants’ informed consent to complete the study protocol, which included repeated saliva collections, use of wearable devices, and ecological momentary assessments of mood, behavior, and physical activity. All procedures were approved by the New York State Institute Institutional Review Board (IRB No. 8149).

**Table 1.** Characteristics of the study cohort (*N* = 25)

| Variable | *M* (*SD*) or count (%) | *Mdn* (*IQR*) | Range |
| --- | --- | --- | --- |
| **Sociodemography** |  |  |  |
| Age [years]<sup>a</sup> | 31.1 (8.0) | 28.5 (13.25) | [21, 47] |
| Sex, assigned | Female: 13 (52%)<br>Male: 12 (48%) |  |  |
| Gender | Woman: 13 (52%)<br>Man: 10 (40%)<br>Diverse: 2 (8%) |  |  |
| Ethnic identity | African American/Black: 6 (24%)<br>Asian: 6 (24%)<br>Hispanic/Latino: 7 (28%)<br>Mixed: 1 (4%)<br>White: 5 (20%) |  |  |
| MacArthur Ladder | 5.8 (2.0) | 6.0 (2.0) | [1, 10] |
| **Lifestyle variables** |  |  |  |
| BMI [kg/m<sup>2</sup>] | 23.9 (3.1) | 23.6 (4.8) | [18.5, 29.5] |
| Daily Activity | 2.4 (1.2) | 2.0 (1.0) | [0, 5] |
| Sleeping quality (PSQI) | 27.6 (12.4) | 24.0 (13.0) | [12, 57] |
| Morningness (MEQ) | 49.9 (6.7) | 49.9 (10.6) | [39.3, 60.5] |
| Daily smoking | 1 (4%) |  |  |
| Alcohol consume | Never: 10 (40%)<br>1-2/week: 7 (28%)<br>1-2/month: 8 (32%) |  |  |
| **Menstrual cycle status<sup>b</sup>** |  |  |  |
| At Sampling Day A | Menstrual: 0 (0%)<br>Follicular: 1 (4%)<br>Early luteal: 3 (12%)<br>Later luteal: 9 (36%)<br>None/Male: 12 (48%) |  |  |
| At Sampling Day B | Menstrual: 2 (8%)<br>Follicular: 1 (4%)<br>Early luteal: 0 (0%)<br>Later luteal: 10 (40%)<br>None/Male: 12 (48%) |  |  |
| **Stress and mood perception** |  |  |  |
| State Anxiety (STAI-S) | 26.0 (10.4) | 23.0 (17.0) | [14, 49] |
| Negative emotional states (mDES) | 6.3 (6.1) | 5.0 (7.0) | [0, 24] |
| Positive emotional states (mDES) | 21.4 (9.1) | 19.0 (12.0) | [1, 35] |
| Daily Hassles [per week] (DHL) | 26.4 (15.8) | 25.0 (26.0) | [0, 55] |
| Social support from friends (PSS-Fr) | 41.0 (8.3) | 39.0 (10.0) | [28, 59] |
| Trait Anxiety (STAI-T) | 32.0 (10.9) | 32.0 (15.0) | [17, 62] |
| Physical fatigue (SFS) | 12.0 (7.6) | 10.0 (11.0) | [0, 26] |
| Mental fatigue (SFS) | 10.9 (8.9) | 9.0 (11.0) | [0, 29] |
| Perceived Helplessness (PSS-10) | 9.2 (5.6) | 8.0 (5.0) | [0, 24] |
| Lack of Self-esteem (PSS-10) | 10.4 (3.4) | 10.0 (5.0) | [5, 16] |
| Depressive symptoms (BDI-II) | 7.9 (10.9) | 4.0 (9.0) | [0, 51] |
| Self-perception of Aging (SPA) | 13.8 (3.6) | 14.0 (6.0) | [6, 18] |

*Note.* <sup>a</sup> One missing value. <sup>b</sup> Cycle phases were estimated using the reported first day of the last menstrual period and typical bleeding duration, assuming a 28-day cycle. Phases were defined as: menstrual (days 1–bleeding duration), follicular (post-menstrual to day 13), ovulatory (days 14–15), early luteal (days 16–21), and late luteal (days 22–28).

### Study Procedures

#### Recruitment, consent, and training

Participants were recruited through flyers posted within CUIMC buildings and through Columbia University’s online recruitment platform, RecruitMe. Interested individuals completed an eligibility screening via REDCap. Eligible participants were contacted by the study coordinator to schedule a video call for informed consent, in which participants provided written informed consent using the REDCap platform. A subsequent video call served to review the study protocol in detail and ensure participants understood the procedures for saliva collection and the completion of surveys. The study coordinator demonstrated each procedure, answered participant questions, and shipped a study kit containing saliva collection tubes and instructional materials to the participants’ home address.

#### Study framework

The study’s data collection period spanned a maximum of 10 days, with the at-home data saliva collection and ecological momentary assessments conducted during day 3 and day 6, with a break of up to 4 days in between. Participants were randomly assigned to one of two timelines, with ‘Timeline 1’ having the first collection day on a weekday and the second collection on a weekend day, vs. ‘Timeline 2’ scheduled vice versa. Participants received a check-in phone call after the first collection to address any concerns and ensure continued compliance.

#### Initial questionnaires

On day 2, the day prior to the first at-home saliva collections, participants completed an initial set of questionnaires, administered through REDCap. The questionnaire battery assessed demographic, health-related, and psychosocial factors, requiring approximately 40−60 minutes to complete. These included: Self-perception of aging [27] and self-rated health [28]; Bem’s Sex-Role Inventory [29]; the MacArthur Ladder [30] to assess the subjective social status; the Stanford Leisure-Time activity categorical (L-Cat) item to assess habitual physical activity [31]; the Pittsburgh Sleep Quality Index (PSQI) [32]; the Morningness and Eveningness Questionnaire (MEQ) [33] to assess circadian rhythm; a 35-item version of the daily hassles list (DHL) [34]; modified Differential Emotions Scale (mDES) to assess the positive and negative affect [35]; Perceived Stress Scale 10-item version (PSS-10) [36] to assess helplessness and lack of self-esteem; Perceived Social Support from Friends (PSS-Fr) [37]; Situation Fatigue Scale (SFS) [38]; State and Trait Anxiety Inventory (STAI-Y1) [39]; Beck Depression Inventory-II (BDI-II) [40].

#### At-home saliva collection

Participants collected saliva in salivettes (Sarstedt #511534500) at multiple timepoints on two separate days (weekday vs. weekend day). On each collection day, saliva samples were taken upon awakening, at 30, 45, and 60 minutes after awakening, and then every hour until bedtime, resulting in a total of up to 20 saliva samples per day (Figure 1A). Time and conditions of saliva collection were recorded using REDCap, a secure web-based platform for data collection and management. Participants completed the REDCap surveys on their mobile device with each sampling to assess their physical activity, and intake of food, beverages, and substances. After completing each saliva collection, participants stored their salivettes in their home freezer. After completing both collection days, they shipped their collected samples back to the laboratory on ice packs. The samples were stored at -80°C before processing to extract cell-free saliva and then stored at -80°C until they were assayed for cell-free DNA and steroid hormone quantification. In total, 828 saliva samples were collected and further processed (see Supplementary Figure S1 for data and specimen availability and timing across sampling time points).

### Saliva processing

Salivettes were thawed and centrifuged at 1,000g × 5 minutes at room temperature. Saliva was collected from the bottom of the outer salivette tubes by pipette. Caution was taken to avoid pipetting any cell pellets that may have precipitated. Saliva was then transferred to 1.5 mL tubes and centrifuged at 5,000g × 10 minutes at room temperature. The resulting supernatants were then transferred to clean 1.5 mL tubes, again taking caution to avoid pipetting precipitates, and stored at -80°C until assay.

### Cell-free mitochondrial DNA assay

Mitochondrial and nuclear cell-free DNA in saliva supernatants were quantified using previously described methods [24] with a few modifications. Briefly, cell-free saliva supernatants were thermolyzed overnight in duplicate. Lysates were analyzed in triplicate using TaqMan chemistry-based real-time quantitative polymerase chain reactions (qPCR) targeting the mitochondrial gene ND1 and the nuclear gene B2M. The medians of triplicate cycle threshold (*C*_T_) values of samples were compared to those of a serial dilution series of a synthetic DNA standard (ranging from 197,542 to 12 copies of ND1 and B2M per reaction) to calculate absolute copy numbers of target genes. The lower limit of detection (LOD) was 24 copies/µl undiluted saliva. Average PCR efficiencies for ND1 and B2M were 91.4% and 91.8%, respectively. The average coefficients of variation of natural log-transformed ND1 and B2M copy number between replicates were 1.2% and 4.7%. Copy numbers were adjusted by correction factors calculated from plate-specific and experiment-wide measurements of reference standards to correct for batch effects. Not detectable B2M values (13.4%) were replaced with the LOD/√2 (see Supplementary Table S2 for details on detection frequencies and missing samples). Additional information about these methods is available in the supplemental material.

### Hormones in saliva

Salivary concentrations of cortisol, cortisone, corticosterone, dehydroepiandrosterone (DHEA), testosterone, progesterone, and estradiol were measured in duplicate for 306 samples using a validated high-throughput liquid chromatography-tandem mass spectrometry (LC-MS/MS) method [41]. The lower limits of detection (LOD) and quantification (LOQ) were as follows: cortisol (LOD = 1.62 pg/ml, LOQ = 5.66 pg/ml), cortisone (LOD = 1.42 pg/ml, LOQ = 4.34 pg/ml), corticosterone (LOD = 1.49 pg/ml, LOQ = 4.90 pg/ml), DHEA (LOD = 10.47 pg/ml, LOQ = 35.55 pg/ml), testosterone (LOD = 2.10 pg/ml, LOQ = 6.86 pg/ml), progesterone (LOD = 1.03 pg/ml, LOQ = 3.38 pg/ml), and estradiol (LOD = 1.26 pg/ml, LOQ = 4.35 pg/ml) [42]. Values below the detection threshold were substituted with the respective LOD/√2. Detailed information on detection frequencies and missing samples for all analytes is provided in Supplementary Table S2.

### Data processing

#### Timing of Samplings

In accordance with established quality control guidelines for cortisol awakening responses [43], we excluded saliva samples that were not time-stamped from the statistical analyses. A tolerance of ±5 minutes was allowed for the first four samples (from awakening to 60 minutes post-awakening), and a tolerance of ±10 minutes was permitted for subsequent samples throughout the day. This resulted in the exclusion of 209 untimed samples, leaving *n* = 619 timed samples for further analysis (see Supplementary Figure S1 for data and specimen availability and timing across sampling time points, and Supplementary Table S3 for frequencies of available biomarker measurements in timed samples). All available time-stamped values were used when calculating associations between biomarkers.

#### Quantification of Diurnal Biomarker Activity

We originally intended to quantify the diurnal dynamics of saliva biomarkers using area under the curve (AUC) measures that integrate all samples collected throughout the day [44–46]. However, the incidence of missing, and especially, mistimed samples resulted in too many missing values (≥48%; Supplementary Figure S1B), limiting the feasibility of this approach. Instead, we characterized diurnal biomarker activity using five complementary metrics, each capturing a distinct dimension of physiological reactivity while reducing missing values: (a) *Awakening level* (S1) — representing the baseline concentration at the start of the day; (b) *Awakening response* (AR) — estimated using the fold change from awakening to the major diurnal peak of the respective biomarker; (c) *Daily average* (AV) — reflecting overall biomarker levels across the day, integrating both stability and fluctuation; (d) *Diurnal variability* (%CV) — the percentage coefficient of variation, indicating how much levels fluctuate relative to the average, thereby quantifying biological instability and reactivity; and (e) *Diurnal AUC above baseline* (AUCi) — estimated using a simplified three-point trapezoidal method based on awakening, peak, and bedtime samples, to approximate the cumulative biomarker output above baseline across the day. Detailed formulas are provided in the supplement. Together, these metrics allow for a multidimensional assessment of biomarker dynamics, capturing not only absolute levels but also the shape, variability, and regulatory characteristics of the diurnal profile. Due to missing and mistimed samplings, the derived metrics could not be computed for a different number of diurnal trajectories (AV: 8%; S1: 18%; %CV: 20%; AR: 38%; AUCi: 44%), depending on the respective formula.

### Statistical analysis

Statistical analyses were conducted using R (version 4.4.2 [47]). Diurnal trajectories were modeled using linear mixed-effects models using the *lme4* package [48]. Outcome variables exhibiting strong right skew were natural log-transformed. All models included random intercepts to account for repeated measures within individuals, nested within day type (weekday vs. weekend) to account for potential differences across sampling days.

Time since awakening was modeled as a fixed effect to estimate mean diurnal trajectories. Moderating effects of covariates (e.g., sex, menstrual cycle phase) were tested via interactions with time. Continuous predictors were mean-centered, and categorical predictors were sum-contrast coded. Post hoc linear contrasts were conducted using the *emmeans* package [49], with *p*-values adjusted for multiple comparisons.

Spearman’s rank correlations (*r*ₛ) were used to examine associations between psychosocial, biological, and lifestyle variables and diurnal biomarker metrics. All statistical tests were two-tailed, with an α level of 0.05 considered statistically significant.

## Results

### Diurnal dynamics and the awakening response of salivary cell-free DNA

Consistent with our prior findings [25], salivary cf-mtDNA exhibited a pronounced awakening response. On average across all participants and days, cf-mtDNA levels rose following awakening, reaching a local peak at 45 minutes (2.5-fold increase, Cohen’s *d* = 0.90, *p*_adj_ = .033) and a diurnal maximum at 180 minutes post-awakening (5.7-fold increase, *d* = 1.70, *p*_adj_ < .001) (Figure 1B). Thereafter, cf-mtDNA levels remained relatively stable, fluctuating between 0.51- and 1.04-fold relative to the 180-minute peak (all *p*_adj_ ≥ .546) (Figure 1C; full post hoc results in Supplementary Data Table D1). Time as a fixed factor explained a small portion of the variance (R²_marg_ = 7.8%), while the full model (R²_cond_ = 58.8%, ICC = 0.553) indicated moderate between-person differences and considerable within-person fluctuations, often diverging from the cohort’s averaged pattern (Figure 1B,C; model details in Supplementary Tables S3, S4).

Blood cf-mtDNA is thought to be released into circulation via both active (e.g., vesicular transport) and passive (e.g., cell death) mechanisms. To assess the contribution of passive DNA release, we also quantified salivary cell-free nuclear DNA (cf-nDNA)—another type of cell-free DNA predominantly released during cell death. Like cf-mtDNA, cf-nDNA showed an awakening response with a diurnal peak at 180 min post-awakening (7.2-fold increase, *d* = 1.60, *p*_adj_ < .001; Figure 1D, cf. Supplementary Figure S2A for additional details), followed by relative stability throughout the day (0.65 to 1.81-fold relative to peak, all *p*_adj_ ≥ .999) (Supplementary Figure S2B; full post hoc results in Supplementary Data Table D1). The average trajectory accounted for 11.8% (R²_marg_) of the variance, with moderate between-person differences and considerable within-person fluctuation across the day (R²_cond_ = 59.0%, ICC = 0.535; model details in Supplementary Tables S3, S4).

Cf-mtDNA and cf-nDNA levels were strongly correlated across all diurnal samples (*r*_S_ = .85, *p* < .001; Figure 2A), with weaker correlations at awakening (S1: *r*_S_ = .49, *p* = .001; Figure 2B) and for the awakening response (AR: *r*_S_ = .47, *p* = .019; Figure 2C). The cf-mtDNA : cf-nDNA ratio significantly decreased across the day (Figure 1D), with post hoc comparisons indicating a progressive decline of up to 2-fold between morning and 240 minutes post-awakening (*p*_adj_ ≤ .026; Supplementary Figure S2C), and relative stability thereafter (Supplementary Figure S2D; full post hoc results in Supplementary Data Table D1). The average trajectory accounted for modest variance (R²_marg_ = 8.0%), with substantial inter- and intra-individual variability in the diurnal change of the cell-free DNA ratio (R²_cond_ = 47.5%, ICC = 0.429; model details in Supplementary Tables S3, S4). These findings suggest a transient decoupling of cf-mtDNA and cf-nDNA dynamics in early hours post-awakening (awakening to ∼180 min), followed by a stable co-regulation later in the day, likely reflecting shared mechanisms of DNA-to-saliva release, such as cell death and turnover.

**Figure 2.**
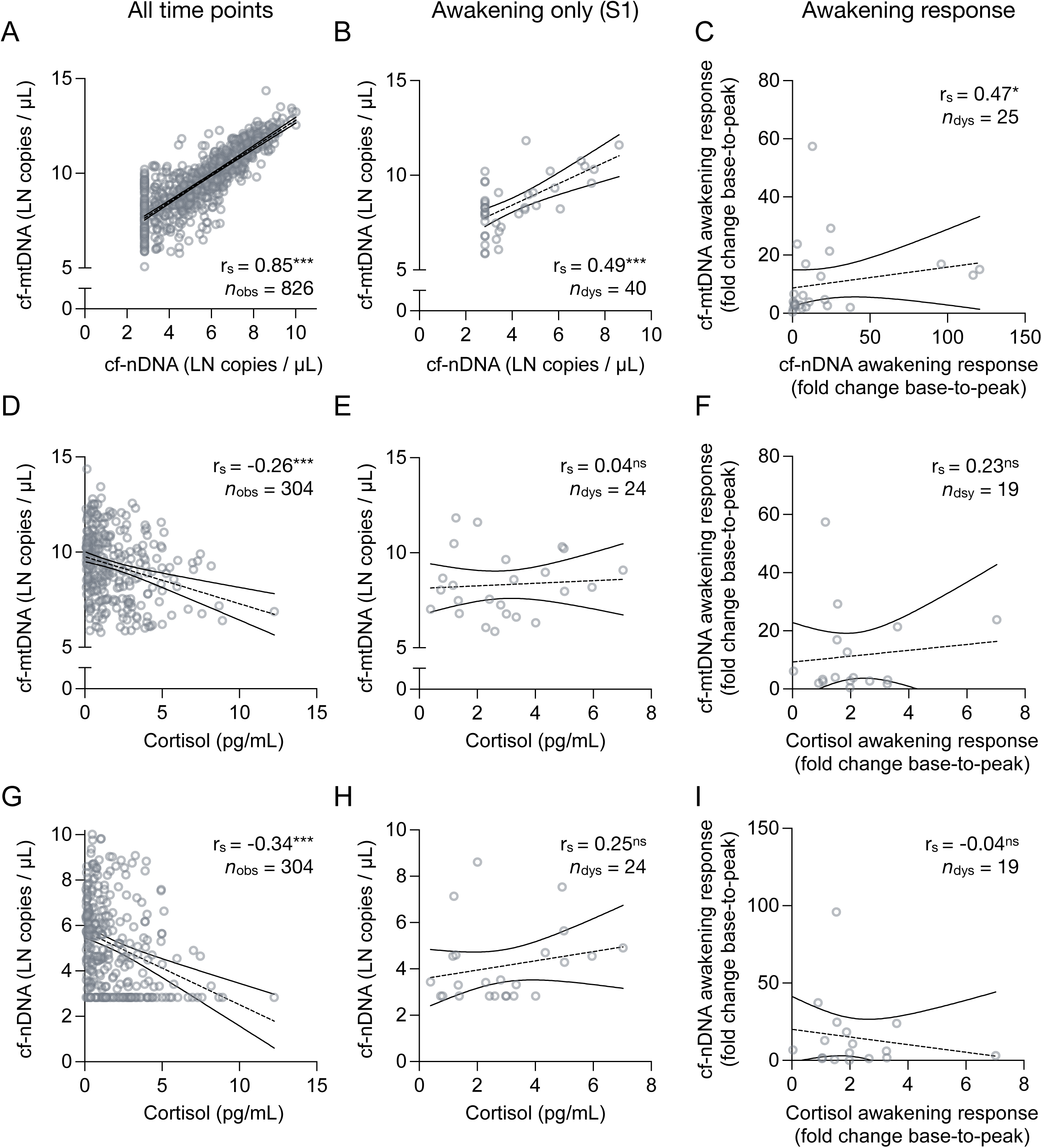
Associations between cell-free DNA and cortisol in saliva. Scatter plots illustrating the correlation between cell-free DNAs and cortisol, with the association of salivary cell-free nuclear DNA (cf-mtDNA) and cell-free mitochondrial DNA (cf-mtDNA) **(A)** over all time points, **(B)** at awakening, and **(C)** awakening responses (fold change from awakening to +180 minutes). Same **(D-F)** for the association of saliva cf-mtDNA and cortisol **(D)** over all time points, **(E)** at awakening, and **(F)** awakening response (cortisol awakening response computed as fold change from awakening to the peak at +30 or +45 min), and **(G-I)** the association of saliva cf-nDNA and cortisol **(G)** over all time points, **(H)** at awakening, and **(I)** awakening response. Linear trend lines and confidence intervals serve illustrative purposes. Effect sizes and two-tailed *p*-values from (A-I) Spearman rank correlations, ^ns^ *p* ≥ 0.05, * *p* < 0.05, ** *p* < 0.01, *** *p* < 0.001.

Cell-free DNA dynamics could not be modelled more accurately when distinguishing weekday and weekend samples as fixed main effect and interaction with time (*p*’s ≥ .495, η²_p_ ≤ .04; Supplementary Table S2; Supplementary Figures S3A). Consistent with our pilot data, we observed moderate within-subject correlations in cf-mtDNA and cf-nDNA concentrations across days (cf-mtDNA: whole-day *r* = .55, *p* < .001; at awakening *r* = .51, *p* = .039; cf-nDNA: whole-day *r* = .53, *p* < .001; at awakening *r* = .61, *p* = .009; Supplementary Figure S3B−E). These findings indicate a degree of within-individual stability in cell-free DNA levels, while also reflecting temporal fluctuations driven by daily physiological processes.

### Cf-mtDNA and cortisol

Our prior report suggested a moderate positive association between the trajectories of saliva cf-mtDNA and cortisol [25]. In the current study, cortisol displayed its typical diurnal pattern, peaking approximately 30 minutes after awakening, followed by a gradual decline throughout the day (Supplementary Figure S4A; model details in Supplementary Tables S3, S4). In line with this, the dynamics of cf-mtDNA and cortisol in saliva showed only modest correlation across matched time points (Figure 2A). While there was a negative correlation between contemporaneous cortisol and cf-mtDNA levels over the entire day (*r*_S_ = −0.26, *p* < .001, Figure 2D), no significant correlation was found in the awakening sample (S1: *r*_S_ = 0.04, *p* = .866, Figure 2E), nor in the awakening response (AR: *r*_S_ = 0.23, *p* = .340, Figure 2F). A similar pattern was observed for cf-nDNA (whole day: *r*_S_ = -0.34, *p* < .001, Figure 2G; S1: *r*_S_ = 0.25, *p* = .242, Figure 2H; AR: *r*_S_ = - 0.04, *p* = .836, Figure 2I). Importantly, the reliability of these statistical associations may be limited by the high rate of missing cortisol data (∼70% across all samples and individuals; 38% of AR values, see Supplementary Tables S2-S3).

Additionally, we measured the diurnal concentrations of other hormones in saliva, including cortisone, corticosterone, testosterone, estradiol, progesterone, and DHEA (with ≥70% missing values as detailed in Supplementary Tables S2-S3). Cortisone, testosterone, and DHEA concentrations exhibited significant changes across the day (Supplementary Table S4-S5; Supplementary Figures S4A-E). We found negative associations of cell-free DNA levels with the levels of cortisone and corticosterone, resembling the results for cortisol. In addition, cf-mtDNA was positively associated with testosterone levels and weakly negatively associated with progesterone and estradiol levels across all available samplings (Supplementary Table S5).

### Psychosocial influences on cell-free DNA in saliva

Similar to other neuroendocrine stress biomarkers, we recently showed that acute psychological stress increases saliva cf-mtDNA levels [10]. Here, we extended these results by investigating the relationship between psychosocial stress and diurnal cell-free DNA dynamics. We investigated the relationships between current mood states (e.g., negative emotions, state anxiety), stress exposure (e.g., 2-week daily hassles), and integrated measures of stress perception and mood disorder symptoms (e.g., trait anxiety, depression) and several metrics to represent complementary aspects of the diurnal cell-free DNA dynamics (full correlation results in Supplementary Data Table D2).

As shown in Figure 3A, our findings reveal that higher awakening levels (S1), a blunted awakening response (AR), and lower diurnal variability (%CV) of cf-mtDNA were each significantly associated with indicators reflecting greater psychosocial stress, including negative emotional states, anxiety, fatigue, depressive symptoms, and lower perceived social support.

**Figure 3.**
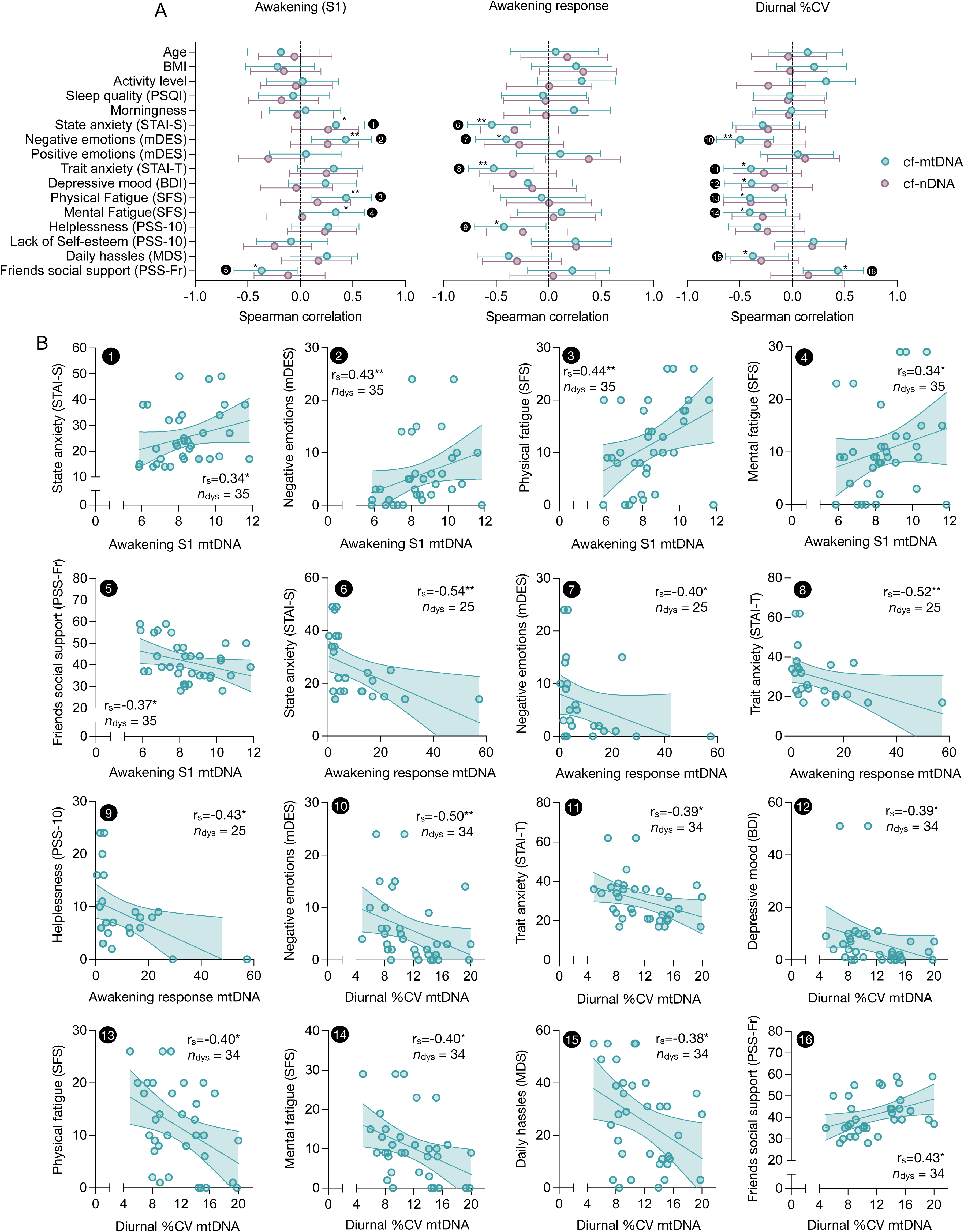
Correlations of saliva cell-free DNA measurements with psychobiological parameters. **(A)** Forest plots illustrating associations between participants’ cell-free mitochondrial DNA (cf-mtDNA, green) and cell-free nuclear DNA (cf-nDNA, purple) diurnal metrics (value at awakening, S1; awakening response (fold change from awakening to +180 minutes); and diurnal percent coefficient of variance, %CV) and psychobiological characteristics. Dots and error bars indicate effect size (as Spearman’s ρ) and 95% confidence interval. Encircled numbers indicate significant correlations illustrated as **(B)** scatter plots. Linear trend lines and 95% confidence intervals serve illustrative purposes. Effect sizes and two-tailed *p-*values from Spearman rank correlations, * *p* < 0.05, ** *p* < 0.01, *** *p* < 0.001.

Specifically, higher awakening levels (S1) of cf-mtDNA were observed among individuals reporting elevated state anxiety (STAI-S: *r*_S_ = .34, *p* = .045), more negative emotional states (mDES: *r*_S_ = .43, *p* = .009), increased physical and mental fatigue (SFS physical: *r*_S_ = .44, *p* = .009; SFS mental: *r*_S_ = .34, *p* = .047), and lower perceived social support from friends (PSS-Fr: *r*_S_ = -.37, *p* = .030).

Conversely, diminished awakening responses of cf-mtDNA (i.e., lower rise from awakening to diurnal peak at 180 min) were associated with elevated state anxiety (STAI-S: *r*_S_ = -.54, *p* = .005), negative emotional states (mDES: *r*_S_ = -.40, *p* = .046), higher trait anxiety (STAI-T: *r*_S_ = -.52, *p* = .007), and more perceived helplessness (PSS-10: *r*_S_ = -.43, *p* = .034).

Following a similar pattern, the diurnal variability (%CV) of cf-mtDNA was lower in individuals reporting more negative emotional states (mDES: *r*_S_ = -.50, *p* = .003), higher trait anxiety (STAI-T: *r*_S_ = -.39, *p* = .021), more (subclinical) depressive symptoms (BDI: *r*_S_ = -.39, *p* = .023), more perceived helplessness (PSS-10: *r*_S_ = -.33, *p* = .055), more fatigue symptoms (SFS physical: *r*_S_ = -.40, *p* = .018; SFS mental: *r*_S_ = -.41, *p* = .017), greater exposure to daily hassles (DHL: *r*_S_ = -.38, *p* = .028), and lower social support from friends (PSS-F: *r*_S_ = .43, *p* = .010) (Figure 3B).

A similar association pattern was present for cf-nDNA (Figure 3A), although the associations were mostly not statistically significant.

We also investigated the relationship between the parameters of diurnal dynamics of cell-free DNAs and biological and lifestyle factors, including age, BMI, assigned sex, social gender, alcohol consumption, chronotype, and habitual sleep quality. We found no significant statistical association between these factors and the diurnal dynamics of cell-free DNAs in saliva. However, there was a trend for a greater diurnal variability (%CV: *r*_S_ = .32, *p* = .064), higher mean concentrations (AV: *r*_S_ = .31, *p* = .051), and higher cumulative output over baseline (AUCi: *r*_S_ = .54, *p* = .009) of cf-mtDNA among individuals reporting more habitual leisure-time activity (Figure 3A, Supplementary Figure S5A, B).

## Discussion

This study provides a detailed characterization of the diurnal saliva cf-mtDNA dynamics under naturalistic conditions over two separate days in healthy individuals. Our key findings demonstrate that saliva cf-mtDNA exhibits a robust awakening response, characterized by a diurnal peak around 180 minutes post-awakening. This pattern suggests a biological rhythmicity, which corroborates and extends the results of our initial study on salivary cf-mtDNA dynamics [25]. In line with this previous report, cf-mtDNA and cf-nDNA were positively correlated across the day, indicating potentially shared release mechanisms. Cell-free DNA dynamics showed limited correspondence with salivary cortisol and other steroid hormones, suggesting asynchronous regulatory mechanisms. Importantly, greater psychological stress experiences, such as daily hassles, negative emotional states, fatigue, and depressive symptoms, were associated with higher awakening cf-mtDNA levels, diminished awakening response, and lower diurnal variability, pointing to an interaction between psychological experiences and mitochondrial signaling in saliva.

The regulation of cf-mtDNA and cf-nDNA release in biofluids remains poorly understood, particularly regarding the influence of psychological and behavioral factors. Previous research has predominantly focused on the acute reactivity of cf-mtDNA in blood and saliva in response to psychological and physiological stress paradigms [10–13, 19]. Our study extends this literature by showing that psychological stressors and related mental states, including daily hassles, reduced social support, negative emotional states, fatigue, and depressive symptoms are associated with basal cf-mtDNA rhythms, with elevated levels at awakening exhibiting the strongest psychobiological associations. At the same time, the diurnal variability of cf-mtDNA was decreased in individuals reporting higher levels of psychosocial stress, an effect that could partly be attributed to higher awakening values.

One unresolved question in the field of cell-free DNA is the extent to which cell-free DNA reflects active secretion (e.g., via vesicular pathways) versus passive release through cell death [20, 21]. In saliva, we observed a tight correlation between cf-mtDNA and cf-nDNA throughout the day, suggesting general co-regulation. However, this coupling temporarily dissipated during the first three hours after awakening, with cf-mtDNA showing a disproportionately higher abundance in saliva than cf-nDNA. This transient decoupling may reflect time-dependent upregulation or enhanced release of mitochondrial DNA specifically, pointing to a potentially distinct post-awakening mechanism. Later in the day, the relatively constant cell-free DNA ratio supports the notion of stable co-release of both nDNA and mtDNA via shared passive pathways (e.g., epithelial turnover, cell rupture) and/or stable semi-regulated mtDNA release (e.g., secretion of exosomes, microvesicles, or entire mitochondria) [20, 21]. Notably, electron microscopy has shown that cf-mtDNA in different biofluids, including saliva, exists in membrane-bound vesicular forms, potentially containing whole mitochondria [10], but more work is required to examine the specific nature of mtDNA-containing structures in saliva. Together, these findings of early-day decoupling and later-day co-regulation may reflect temporal shifts in the cellular origin, release mechanism, or regulatory control of cf-mtDNA versus cf-nDNA.

This raises a broader question: what is the biological function of releasing cf-mtDNA into saliva, particularly in response to stress exposure? Cf-mtDNA release into the circulation has been proposed to serve various functions, including modulation of inflammatory responses and intercellular signaling [9, 21]. In saliva, the observed diurnal rhythms, sensitivity to psychosocial stress, and transient decoupling from cf-nDNA dynamics suggest at least two functional roles for cf-mtDNA: one reflecting basal physiological turnover under endogenous control—possibly linked to circadian and/or behavioral cues—and another reflecting acute physiological adaptation to early-day activation and psychosocial or physical stress exposure. Although speculative, the transient surge in salivary cf-mtDNA content may be part of an anticipatory response that facilitates tissue repair during stressful situations. Notably, application of mitochondria-rich platelets to injured tissue has been shown to promote wound healing in mouse models, where the release and uptake of free mitochondria by stromal cells was necessary [50]. This represents a form of cell-to-cell mitochondrial transfer with reparative effects [51]. In this context, we speculate that the act of licking wounds could apply mitochondria-rich saliva—including cf-mtDNA—to injured tissue, potentially stimulating local repair mechanisms such as immune activation and epithelial regeneration. While direct evidence in humans is lacking, it is conceivable that stress-induced elevations in saliva cf-mtDNA reflect an evolutionarily conserved mechanism to enhance tissue readiness and repair capacity in anticipation of injury. The post-awakening rise in saliva cf-mtDNA might accordingly reflect a biological shift toward increased readiness for environmental engagement and potential stress exposure.

Several key questions remain to be resolved to substantiate such considerations. To advance this line of research, we outline several key questions and propose directions for future investigation: First, what are the precise cellular sources and release pathways for cf-mtDNA into saliva? Advanced proteomic and vesicle-isolation studies are needed to identify whether these fragments stem from epithelial cells, immune cells, or filtered plasma. Second, it remains unclear whether the cf-mtDNA in saliva possesses signaling properties or is merely a byproduct of cell turnover. The application of mitochondrial membrane-specific dyes, electron microscopy, and single-vesicle profiling may help address this question [e.g., 10]. Third, the potential for cf-mtDNA to serve as a stress-related biomarker in mental health research warrants further investigation. Our findings highlight that diurnal variability of cf-mtDNA is sensitive to trait anxiety, depressive symptoms, and daily stressors, opening up the possibility of using this metric in ecologically valid, repeated-measure assessments of stress vulnerability or resilience. Lastly, given the non-invasive nature of saliva sampling, cf-mtDNA may be a feasible tool for large-scale or vulnerable population studies where blood collection is impractical.

### Limitations

Despite its contributions, our study has limitations. The sample size was modest but consistent with previous intensive-sampling protocols and in line with recommendations for covering weekday-weekend differences [43]. Although we observed moderate to strong within-day and within-subject consistency, including additional sampling days would allow for more precise assessment of test–retest reliability of cell-free DNA dynamics and conclusions about trait-like individual differences [43, 44]. Missing data, particularly for steroid hormone quantification (∼70% missing), limited our ability to fully evaluate hormonal co-dynamics of cell-free DNA. However, previous studies could not demonstrate a contemporaneous coupling of steroid and cell-free DNA release in saliva, at least upon acute stress exposure [13]. Lastly, sleep, activity/exercise, and diurnal diet patterns are likely to modulate cell-free DNA dynamics, calling forth the need for additional investigations using tracking device data and experimental trials [52]. The study cohort consisted of healthy, predominantly non-smoking, middle-aged adults, highlighting the need for further research to generalize the findings to other age groups and clinical populations.

### Conclusions

This study expands our understanding of cf-mtDNA dynamics in human saliva. By establishing a reproducible diurnal rhythm, a robust awakening response, and associations with self-reported measures of psychosocial stress, our findings provide the foundation for future work exploring the psychobiological regulation of salivary cf-mtDNA. Beyond its mechanistic interest, cf-mtDNA emerges as a promising, non-invasive marker of mitochondrial stress signaling, with potential applications in mental health and behavioral medicine. Moving forward, studies integrating physiological monitoring, broader biomarker panels, and mechanistic cellular assays will be needed to clarify the biological role and diagnostic utility of saliva cf-mtDNA in human health.

## Supporting information

Supplementary Data Table D1 (post hoc tests)

Supplementary Data Table D2 (correlation analyses)

## Data Availability

All data produced in the present study are available upon reasonable request to the authors

## Acknowledgements

This work was supported by the National Institutes of Health (R21MH123927) awarded to M.P., by a Postdoctoral Research Fellowship from the Fulbright Commission awarded to A.B., and a Fellowship of the G.A. Lienert Foundation awarded to A.B..

## Author contributions

C.T. and M.P. designed the study, with A.G. assisting in organizing the study setup and obtaining legal and ethical approvals. S.L. coordinated data and sample collection from participants. D.S. and A.P. conducted the cell-free DNA assays. C.K. provided the methodology for steroid hormone measurement. A.B. performed the statistical analyses and drafted the manuscript with support from C.T. and D.S. All authors reviewed and approved the final version of the manuscript.

## Supplementary information

is available at MP’s website.

