## Supplementary Materials (methods, tables) for "Diurnal dynamics and psychobiological regulation of cell-free mitochondrial and nuclear DNA in human saliva"

| | |
|---|---|
| Alexander Behnke, PhD | Caroline Trumpff, PhD |
| Universität Ulm | Columbia University Irving Medical Center |
| Albert-Einstein-Allee 47 | 622 West 168th St., PH 1540O |
| D-89081 Ulm, Germany | New York, NY 10032 |
| Tel.: +49 731 5026597 | Tel: +1 703 8701609 |
| | |

### Supplementary Methods

#### *Inclusion and exclusion criteria*

Inclusion Criteria:

- Healthy male and female adults aged 20−50 years.
- Ability and willingness to provide saliva samples, wear actigraphy and heart rate monitors, and complete questionnaires.
- Capacity to provide informed consent and participate in the study.
- For female participants, use of an effective birth control method.
- Body Mass Index (BMI) between 18.5 and 29.9, calculated from self-reported height and weight.
- Fluency in English.
- Regular menstrual cycles (21−35 days) for premenopausal women, specifically during the luteal phase (days 21−28 of the cycle).
- Access to a freezer for saliva sample storage.

Exclusion Criteria:

- History or current diagnosis of oral health issues (e.g., periodontitis, gingivitis).
- Diagnosis of early menopause (absence of menses for ≥2 months).
- Employment as a shift worker.
- Diagnosis of sleep disorders, including insomnia.
- Current pregnancy or potential pregnancy.
- Active cancer or chronic inflammatory disorders.
- Use of medications affecting cardiovascular or inflammatory systems (e.g., NSAIDs, corticosteroids, β-blockers).
- Presence of cold or flu symptoms within the last four weeks.
- Current use of nicotine or tobacco products.

#### *Cell-free mitochondrial DNA quantification*

Mitochondrial and nuclear DNA in cell-free saliva were quantified using a previously established method [1] with the following changes and additional details. In instances where the cited method indicates that plates are to be vortexed, they were instead shaken at 1,500 rpm for 1 minute on an OrbiShaker MP (Benchmark Scientific, Sayreville, NJ) and then briefly centrifuged using an Axygen plate centrifuge (Corning Life Sciences, Corning, NY). Models of thermal cyclers used for lysis were either T100 (Bio-Rad, Hercules, CA) or ProFlex PCR System (Applied Biosystems, Waltham, MA). Instead of preparing standard curves using DNA extracted from human fibroblasts (hFB), they were prepared using a chimeric ND1-B2M synthetic DNA fragment (sND1-B2M; gBlocks® gene fragment, Integrated DNA Technologies, Coralville, IA). Both templates are described in the published method [1]. The absolute copy numbers of target genes in sND1-B2M were determined by quantifying it against hFB, which had been previously quantified by digital droplet polymerase chain reaction (Figure SX1). Instead of using the averages of triplicate $C_T$ measurements after discarding outliers for quantification, the medians of triplicate $C_T$ measurements ($C_{Tmed}$) without outlier removal were used. For each qPCR plate, linear regressions were fit to $C_{Tmed}$ versus natural log-transformed (Ln) copy number for each target gene in the serially diluted standard. Absolute copy numbers of target genes in samples were calculated using the following formula:

$$Copy\ per\ reaction = e^{\frac{C_{Tmed}-b}{m}}$$

Where $b$ and $m$ are the slope and intercept of the linear regression of the standard curve for the corresponding qPCR plate and target gene. A target gene was considered below the limit of detection (LOD) when its copy number per reaction was three or fewer [2]. Copy per reaction values were transformed to copy per µL saliva by correcting for dilution of samples into lysis and qPCR buffers. Eight reference standards, created by pooling samples from the Picard lab biobank, were loaded on each 96-well plate alongside saliva samples prior to lysis to assess and correct for plate effects. The measurements of these standards were used to calculate correction factors that were specific to each pair of replicate plates as follows:

$$cf_p = \frac{\overline{LN(ND1)}_p}{\overline{LN(ND1)}_{exp}}$$

Where $cf_p$ is the correction factor for plate $p$, and $\overline{LN(ND1)}_p$ and $\overline{LN(ND1)}_{exp}$ are the averages of LN ND1 copy number per µL in reference standards on plate $p$ and experiment-wide, respectively. All measurements of ND1 and B2M in samples were then adjusted by dividing their LN values by the correction factor for their corresponding plate. Adjusted values were used in subsequent statistical analyses.

#### *Diurnal Biomarker Metrics*

We computed five different metrics to capture complementary aspects of the shape, variability, and regulatory characteristics of the diurnal biomarker profile. For the calculation, we used the following formula, with $y_i$ indicating the values of respective biomarker at time $t_i$ indicating the effective sampling time point, ranging from Awakening ($i$ = 1) to a maximum of $i_{max}$ = 20 samplings per day:

(a) Awakening level: S1 = $\ln(y_1)$

(b) Awakening response: AR = $\frac{y_{peak}}{y_1}$ , using the concentrations at the diurnal peak ($y_{peak}$) located at $t$ = 180 min for cell-free DNAs and at $t$ = 30 to 45 min for cortisol.

(c) Daily average: AV = $Mean(\ln(y_i))$, if at least 60% of the samplings per day were available and timed

(d) Diurnal variability: %CV = $\frac{SD(\ln(y_i))}{Mean(\ln(y_i))} \cdot 100\%$

(e) Three-point diurnal AUC above baseline: AUCi = $\sum_{i=1}^{n-1}\left(\frac{y_i+y_{i+1}}{2} \cdot (t_{i+1} - t_1)\right) - (t_n - t_1) \cdot y_1$, where $y_i$ are the $\log_e$-scaled values of respective biomarker at the sampling time points $t_i$, with $t_1$ = Awakening, $t_2$ = Peak, $t_3$ = Bedtime. Computation is only done if all $n$ = 3 time points are available. Note that $t_2$ was +180 min post-awakening for cell-free DNAs and the maximum value at either +30 or +45 min post-awakening for cortisol.

Note that simplified estimations were used, for example, for the awakening response, as the amount of missing and not-timed samplings did not allow to compute areas under the curve integrating all samplings from awakening to the marker's diurnal peak [3].

### Supplementary Tables

**Supplementary Table S1**. Detection and missing frequencies of salivary biomarkers among all available samples (*n* = 828)

| | Quantifiably detected [a] | Semi-quantifiably detected [b] | Not detected [c] | Not measured |
|---|---|---|---|---|
| Cf-mtDNA | 826 (99.8%) | – | – | 2 (0.2%) |
| Cf-nDNA | 715 (86.4%) | – | 111 (13.4%) | 2 (0.2%) |
| Cortisol | 306 (37.0%) | – | – | 520 (62.8%)[d] |
| Cortisone | 306 (37.0%) | – | – | 520 (62.8%)[d] |
| Corticosterone | 96 (11.6%) | – | 210 (25.4%) | 520 (62.8%)[d] |
| DHEA | 204 (24.6%) | – | 102 (12.3%) | 520 (62.8%)[d] |
| Testosterone | 262 (31.6%) | 44 (5.3%) | – | 520 (62.8%)[d] |
| Progesterone | 306 (37.0%) | – | – | 520 (62.8%)[d] |
| Estradiol | 154 (18.6%) | 108 (13.0%) | 44 (5.3%) | 520 (62.8%)[d] |

*Note*: Absolute frequencies and percentages relative to the total number of available saliva samples, *n* = 828, collected from 25 participants over two days, with a maximum of 20 samples per day. [a] Concentrations were above the limit of quantification (LOQ). [b] Concentrations were below the limit of quantification (LOQ) and above the limit of detection (LOD). [c] Concentrations not detectable or below the LOD were replaced with the LOD/√2. [d] The measurement of cell-free DNA was prioritized over hormone measurements in terms of available material and financial resources, ultimately resulting in 306 saliva samples being used for hormone measurements.

**Supplementary Table S2**. Detection and missing frequencies of salivary biomarkers among timed samples ($n$ = 576)

| | Quantifiably detected [a] | Semi-quantifiably detected [b] | Not detected [c] | Not measured |
|---|---|---|---|---|
| Cf-mtDNA | 574 (99.7%) | − | − | 2 (0.3%) |
| Cf-nDNA | 489 (84.9%) | − | 85 (14.8%) | 2 (0.3%) |
| Cortisol | 247 (42.9%) | − | − | 329 (57.1%) |
| Cortisone | 247 (42.9%) | − | − | 329 (57.1%) |
| Corticosterone | 83 (14.4%) | − | 164 (28.5%) | 329 (57.1%) |
| DHEA | 168 (29.2%) | − | 79 (13.7%) | 329 (57.1%) |
| Testosterone | 219 (38.0%) | 28 (4.9%) | − | 329 (57.1%) |
| Progesterone | 247 (42.9%) | − | − | 329 (57.1%) |
| Estradiol | 120 (20.8%) | 87 (15.1%) | 40 (6.9%) | 329 (57.1%) |

*Note*: Absolute frequencies and percentages relative to the number of timely collected saliva samples, $n$ = 576, collected from 25 participants over two days, with a maximum of 20 samples per day. The measurement of cell-free DNA was prioritized over hormone measurements in terms of available material and financial resources, ultimately resulting in 306 saliva samples being used for hormone measurements, of which 247 were collected timely. [a] Concentrations were above the limit of quantification (LOQ). [b] Concentrations were below the limit of quantification (LOQ) and above the limit of detection (LOD). [c] Concentrations not detectable or below the LOD were replaced with the LOD/√2.

**Supplementary Table S3**. Results of model comparisons for the relevance of sampling day

| Outcome | Fixed terms | Random terms | AICc | BIC | $R^2_{marg}$ | $R^2_{cond}$ |
|---|---|---|---|---|---|---|
| Cf-mtDNA | Time | ID + ID:Day | **12944.9** | **13043.0** | **.078** | **.588** |
| | Time × Day | ID + ID:Day | 12970.5 | 13150.5 | .090 | .596 |
| Cf-nDNA | Time | ID + ID:Day | **8458.3** | **8583.5** | **.118** | **.590** |
| | Time × Day | ID + ID:Day | 8515.4 | 8695.4 | .128 | .594 |
| DNA ratio | Time | ID + ID:Day | **6105.9** | **6204.0** | **.080** | **.475** |
| | Time × Day | ID + ID:Day | 6125.3 | 6305.4 | .107 | .480 |
| Cortisol | Time | ID + ID:Day | **533.4** | **609.1** | **.585** | **.734** |
| | Time × Day | ID + ID:Day | 566.2 | 698.5 | .593 | .736 |
| Cortisone | Time | ID + ID:Day | **1305.6** | **1381.4** | **.555** | **.675** |
| | Time × Day | ID + ID:Day | 1336.4 | 1468.7 | .565 | .681 |
| Testosterone | Sex × Time | ID + ID:Day | **1844.5** | **1974.2** | **.874** | **.941** |
| | … + Time × Day | ID + ID:Day | 1879.8 | 2054.9 | .872 | .943 |
| Progesterone | SexCycle × Time | ID + ID:Day | **915.0** | **1082.1** | **.162** | **.742** |
| | … + Time × Day | ID + ID:Day | 971.5 | 1171.8 | .175 | .742 |
| DHEA | Time | ID:Day[a] | **2851.6** | **2924.3** | **.148** | **.181** |
| | Time × Day | ID:Day[a] | 2884.9 | 3014.6 | .193 | .229 |
| Estradiol | Sex × Time | ID:Day[a] | **1357.5** | **1484.6** | **.142** | **.162** |
| | … + Time × Day | ID:Day[a] | 1375.1 | 1548.3 | .250 | .310 |

*Note.* Linear mixed-effects models with transformed outcome scales using the natural logarithm to adjust excessive right skew, except for DNA ratio. We modelled a repeated data structure using random intercepts for subjects and sampling day (weekday vs. weekend) nested within subjects. To check whether time trends systematically differ between weekdays and weekend, we considered the sampling day as fixed effect. Model fit indices (AICc, BIC) suggest that sampling day does not improve the model fit when considered as a fixed effect. Thus, all main results are based on models specifying the sampling day as a random intercept only. Models mapping the trajectories of hormones with sex effects and/or effects of the menstrual cycle specified either assigned sex (here, concordant with gender identity) or a combined sex/phase variable (male vs. female-follicular vs. female-luteal) as null model. [a] Random intercept for subjects (ID) was singular and has been removed.

**Supplementary Table S4**. Results of linear mixed-effects models of diurnal biomarker trajectories

| **Outcome** | **Model terms** | **Test statistics** | | | **Effect size** |
|---|---|---|---|---|---|
| Cf-mtDNA | *Fixed effects* | ***F*** | ***df*s** | ***p*** | **$\eta^2_{part}$** |
| | Intercept | 2009.84 | 1, 24.04 | < .001 | |
| | Time | 5.28 | 19, 513.62 | < .001 | .16 |
| | *Random effects* | **LR-$\chi^2$** | ***df*** | ***p*** | **$\sigma_{random}$** |
| | ID | 9.77 | 1 | .002 | 0.93 |
| | ID:Day | 50.68 | 1 | < .001 | 0.67 |
| | *Overall model* | ***F*** | ***df*s** | ***p*** | **$R^2_{marg}$ ($R^2_{cond}$)** |
| | | 5.28 | 19, 513.62 | < .001 | .078 (.588) |
| Cf-nDNA | *Fixed effects* | ***F*** | ***df*s** | ***p*** | **$\eta^2_{part}$** |
| | Intercept | 503.19 | 1, 24.04 | < .001 | |
| | Time | 7.89 | 19, 514.04 | < .001 | .23 |
| | *Random effects* | **LR-$\chi^2$** | ***df*** | ***p*** | **$\sigma_{random}$** |
| | ID | 10.62 | 1 | .001 | 1.09 |
| | ID:Day | 56.34 | 1 | < .001 | 0.76 |
| | *Overall model* | ***F*** | ***df*s** | ***p*** | **$R^2_{marg}$ ($R^2_{cond}$)** |
| | | 7.89 | 19, 514.04 | < .001 | .118 (.590) |
| DNA ratio | *Fixed effects* | ***F*** | ***df*s** | ***p*** | **$\eta^2_{part}$** |
| | Intercept | 870.29 | 1, 24.03 | < .001 | |
| | Time | 4.29 | 19, 518.74 | < .001 | .14 |
| | *Random effects* | **LR-$\chi^2$** | ***df*** | ***p*** | **$\sigma_{random}$** |
| | ID[a] | 21.31 | 1 | < .001 | 0.63 |
| | ID:Day | 6.75 | 1 | .009 | 0.23 |
| | *Overall model* | ***F*** | ***df*s** | ***p*** | **$R^2_{marg}$ ($R^2_{cond}$)** |
| | | 4.29 | 19, 518.74 | < .001 | .080 (.475) |
| Cortisol | *Fixed effects* | ***F*** | ***df*s** | ***p*** | **$\eta^2_{part}$** |
| | Intercept | 3.09 | 1, 18.15 | .096 | |
| | Time | 24.20 | 19, 205.85 | < .001 | .69 |
| | *Random effects* | **LR-$\chi^2$** | ***df*** | ***p*** | **$\sigma_{random}$** |
| | ID | 6.91 | 1 | .009 | 0.41 |
| | ID:Day | 1.14 | 1 | .284 | 0.14 |
| | *Overall model* | ***F*** | ***df*s** | ***p*** | **$R^2_{marg}$ ($R^2_{cond}$)** |
| | | 24.20 | 19, 205.85 | < .001 | .585 (.734) |
| Cortisone | *Fixed effects* | ***F*** | ***df*s** | ***p*** | **$\eta^2_{part}$** |
| | Intercept | 470.47 | 1, 18.10 | < .001 | |
| | Time | 19.34 | 19, 206.15 | < .001 | .64 |

| | | | | | |
|---|---|---|---|---|---|
| | *Random effects* | **LR-$\chi^2$** | ***df*** | ***p*** | **$\sigma_{random}$** |
| | ID | 3.63 | 1 | .057 | 0.24 |
| | ID:Day | 2.49 | 1 | .114 | 0.14 |
| | *Overall model* | ***F*** | ***df*s** | ***p*** | **$R^2_{marg}$ ($R^2_{cond}$)** |
| | | 19.34 | 19, 206.15 | < .001 | .555 (.675) |
| Testosterone | *Fixed effects* | ***F*** | ***df*s** | ***p*** | **$\eta^2_{part}$** |
| | Intercept | 605.07 | 1, 121.31 | < .001 | |
| | Sex | 49.49 | 1, 121.56 | < .001 | .29 |
| | Time | 3.22 | 19, 184.12 | < .001 | .25 |
| | Time × Sex | 1.83 | 18, 184.19 | .024 | .15 |
| | *Random effects* | **LR-$\chi^2$** | ***df*** | ***p*** | **$\sigma_{random}$** |
| | ID | 8.09 | 1 | .004 | 0.24 |
| | ID:Day | 11.26 | 1 | < .001 | 0.14 |
| | *Overall model* | ***F*** | ***df*s** | ***p*** | **$R^2_{marg}$ ($R^2_{cond}$)** |
| | | 9.09 | 38, 173.89 | < .001 | .874 (.941) |
| Progesterone | *Fixed effects* | ***F*** | ***df*s** | ***p*** | **$\eta^2_{part}$** |
| | Intercept | 413.21 | 1, 91.96 | < .001 | |
| | SexCycle | 0.28 | 2, 99.03 | .754 | .00 |
| | Time | 1.26 | 19, 171.58 | .217 | .12 |
| | Time × SexCycle | 1.30 | 33, 169.88 | .146 | .20 |
| | *Random effects* | **LR-$\chi^2$** | ***df*** | ***p*** | **$\sigma_{random}$** |
| | ID | 25.95 | 1 | < .001 | 0.21 |
| | ID:Day | 0.76 | 1 | .382 | 0.03 |
| | *Overall model* | ***F*** | ***df*s** | ***p*** | **$R^2_{marg}$ ($R^2_{cond}$)** |
| | | 1.09 | 54, 157.91 | .343 | .162 (.742) |
| DHEA | *Fixed effects* | ***F*** | ***df*s** | ***p*** | **$\eta^2_{part}$** |
| | Intercept | 1049.97 | 1, 19.90 | < .001 | |
| | Time | 2.18 | 19, 214.04 | .004 | .16 |
| | *Random effects* | **LR-$\chi^2$** | ***df*** | ***p*** | **$\sigma_{random}$** |
| | ID[a] | – | – | – | – |
| | ID:Day | 1.19 | 1 | .275 | 0.27 |
| | *Overall model* | ***F*** | ***df*s** | ***p*** | **$R^2_{marg}$ ($R^2_{cond}$)** |
| | | 2.18 | 19, 214.04 | .004 | .148 (.181) |
| Estradiol | *Fixed effects* | ***F*** | ***df*s** | ***p*** | **$\eta^2_{part}$** |
| | Intercept | 11.41 | 1, 207.92 | < .001 | |
| | Sex | 0.00 | 1, 208.00 | .998 | .00 |
| | Time | 0.89 | 19, 197.43 | .598 | .08 |
| | Time × Sex | 1.11 | 18, 197.50 | .348 | .09 |

| *Random effects* | **LR-$\chi^2$** | ***df*** | ***p*** | **$\sigma_{random}$** |
|---|---|---|---|---|
| ID[a] | – | – | – | – |
| ID:Day | 0.35 | 1 | .555 | 0.14 |
| *Overall model* | ***F*** | ***df*s** | ***p*** | **$R^2_{marg}$ ($R^2_{cond}$)** |
| | 1.07 | 38, 186.69 | .373 | .142 (.162) |

*Note*. Outcomes were transformed using the natural logarithm to adjust excessive right skew. Models mapping the trajectories of hormones with sex effects and/or effects of the menstrual cycle specified either assigned sex (here, concordant with gender identity) or a combined sex/phase variable (male vs. female-follicular vs. female-luteal). [a] Random intercept for subjects (ID) was singular and has been removed. Analyses of cell-free mitochondrial DNA (cf-mtDNA) and cell-free nuclear DNA (cf-nDNA) and their ratio based on $n_{obs}$ = 574 observations in 49 trajectories from $N$ = 25 participants; cortisol, cortisone, testosterone, progesterone, DHEA, and estradiol models based on $n_{obs}$ = 247 observations in 33 trajectories from $N$ = 17 participants. Post hoc tests for significant effects are provided in the Supplementary Data Table D1.

**Supplementary Table S5**. Spearman correlations between cell-free DNA and hormone levels in saliva in contemporaneous samples ($n$ = 304)

| | ***Cf-mtDNA*** | | ***Cf-nDNA*** | |
|---|---|---|---|---|
| | $r_S$ | $p$ | $r_S$ | $p$ |
| Cortisol | −.26 | < .001 | −.34 | < .001 |
| Cortisone | −.25 | < .001 | −.32 | < .001 |
| Corticosterone | −.26 | < .001 | −.21 | < .001 |
| DHEA | −.06 | .284 | −.10 | .087 |
| Testosterone | .27 | < .001 | .07 | .242 |
| Progesterone | −.15 | .008 | .01 | .858 |
| Estradiol | −.13 | .027 | −.09 | .135 |
