## Supplementary Figures for "Diurnal dynamics and psychobiological regulation of cell-free mitochondrial and nuclear DNA in human saliva"

Figure S1

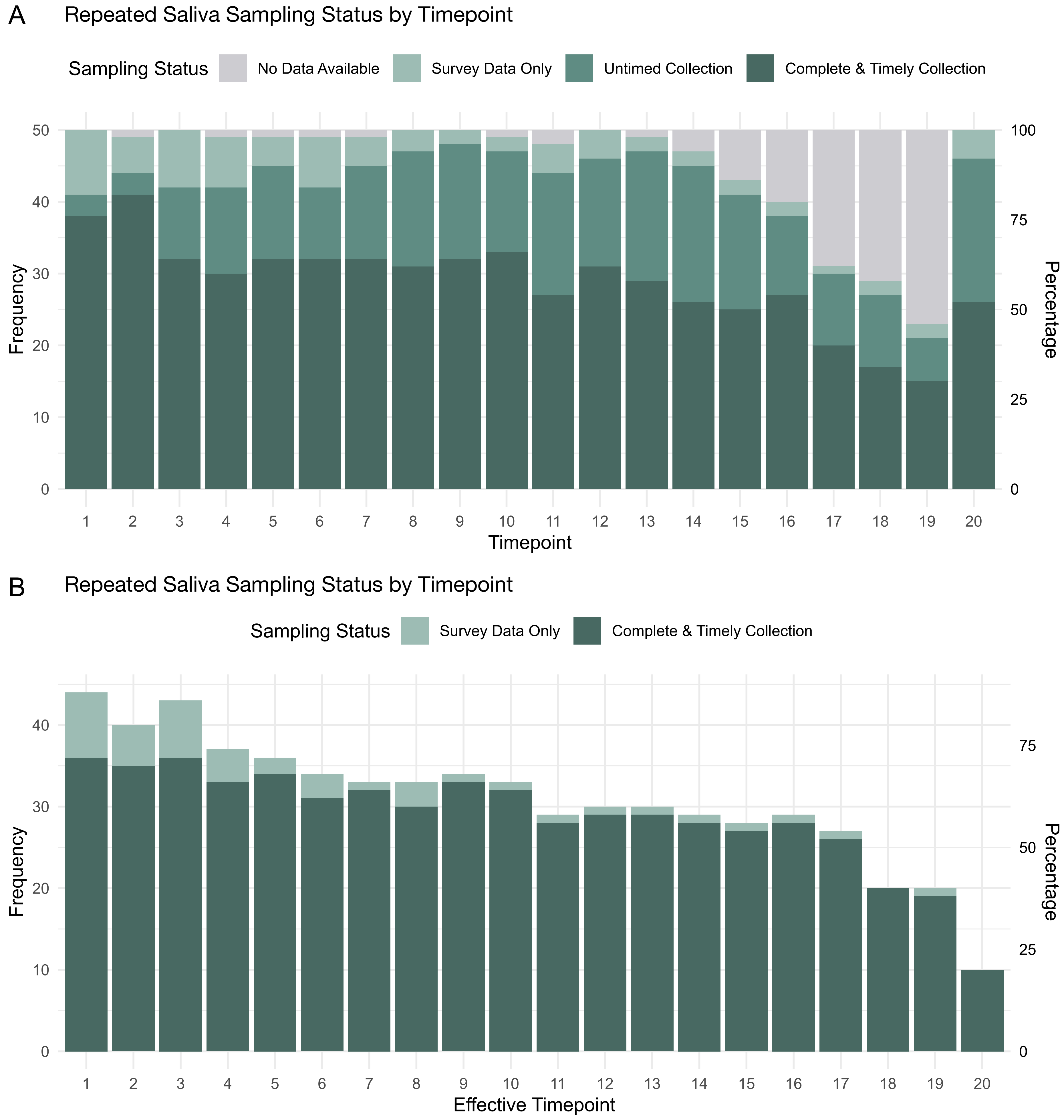


**Supplemental Figure S1. Data and specimen availability by sampling time point. (A)** Available data and saliva specimen collections across sampling time points. Data were collected from 25 participants who responded to a mobile survey and provided saliva samples over two days at up to 20 time points, yielding a maximum of 50 data points and specimen collections per time point. 'Complete & Timely Collection' (dark green) indicates complete and timely saliva collection with a corresponding survey response; 'Survey Data Only' (light green) indicates time points where only survey responses were available, but no saliva was collected; 'Untimed collection' indicates not timed sampling of saliva and survey data. **(B)** Available survey data and saliva collections after quality control for correct timing. The frequency of available data decreases as the day progresses, as participants cease to collect samples when going to bed.

Figure S2

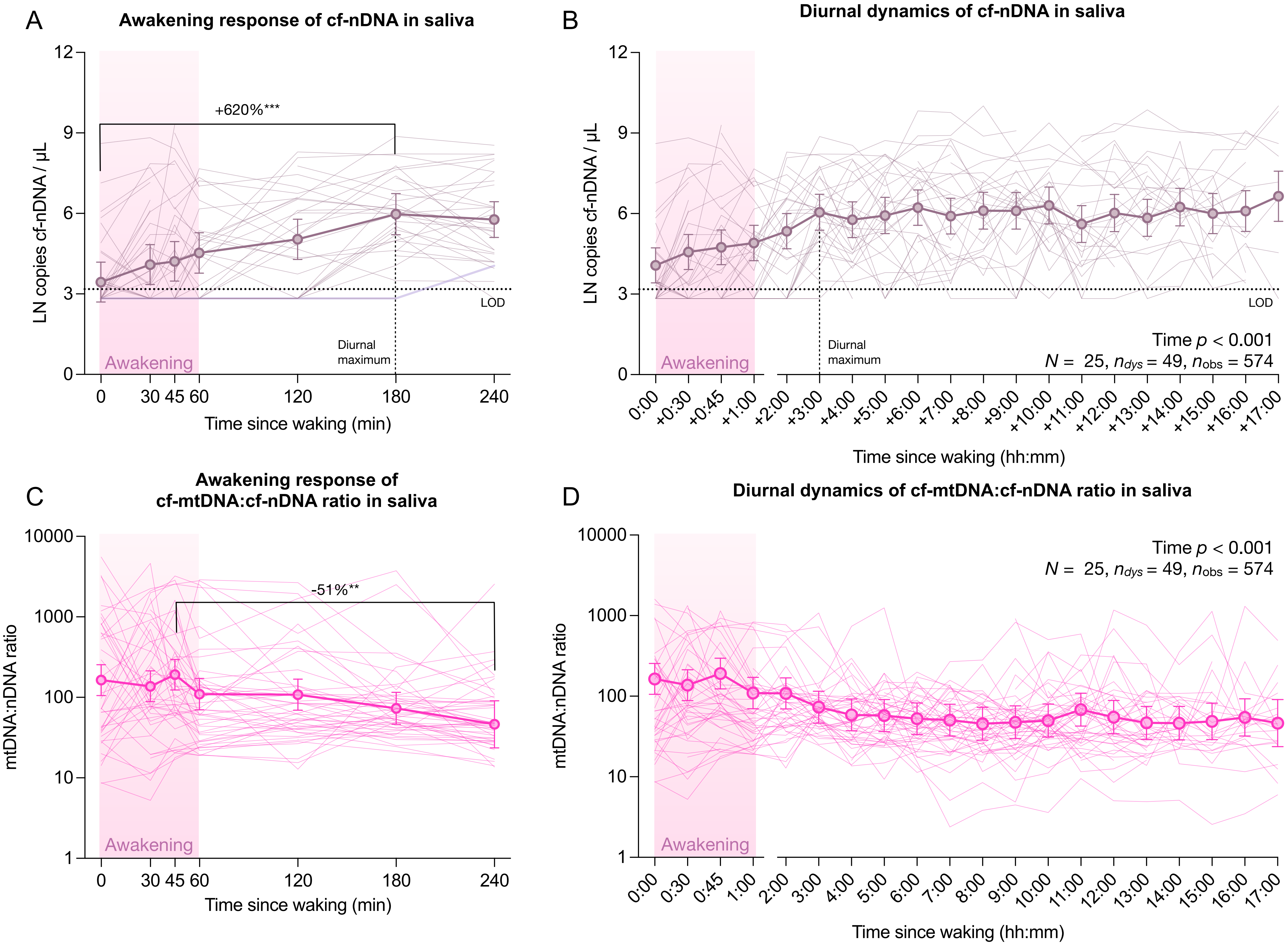


**Supplemental Figure S2. Trajectories of cell-free nuclear DNA and the ratio of cell-free mitochondrial to nuclear DNA in saliva**. Diurnal saliva cell-free nuclear DNA (cf-nDNA) trajectories for **(A)** the awakening response, and **(B)** for all time points. Horizontal dotted line indicates the limit of detection for B2M, the qPCR target representing nDNA; vertical dotted line indicates the time point of the diurnal maximum of the mean cf-nDNA trajectory. **(C,D)** Same as (A,B) for the ratio of cell-free mitochondrial DNA (cf-mtDNA) to cf-nDNA in saliva. Values are presented as (A,B) natural log-transformed concentrations (copies / µl saliva) and (C,D) linear ratios of mtDNA:nDNA. Thin lines represent individual trajectories, dots and error bars indicate linear mixed-effects model-derived marginal means and 95% confidence intervals. Effect sizes and *p*-values from linear mixed-effects models and Tukey's *post hoc* tests, ** $p_{Tukey} < 0.01$, *** $p_{Tukey} < 0.001$, two-tailed. Only the post hoc comparisons that indicate the greatest changes were plotted.

Figure S3

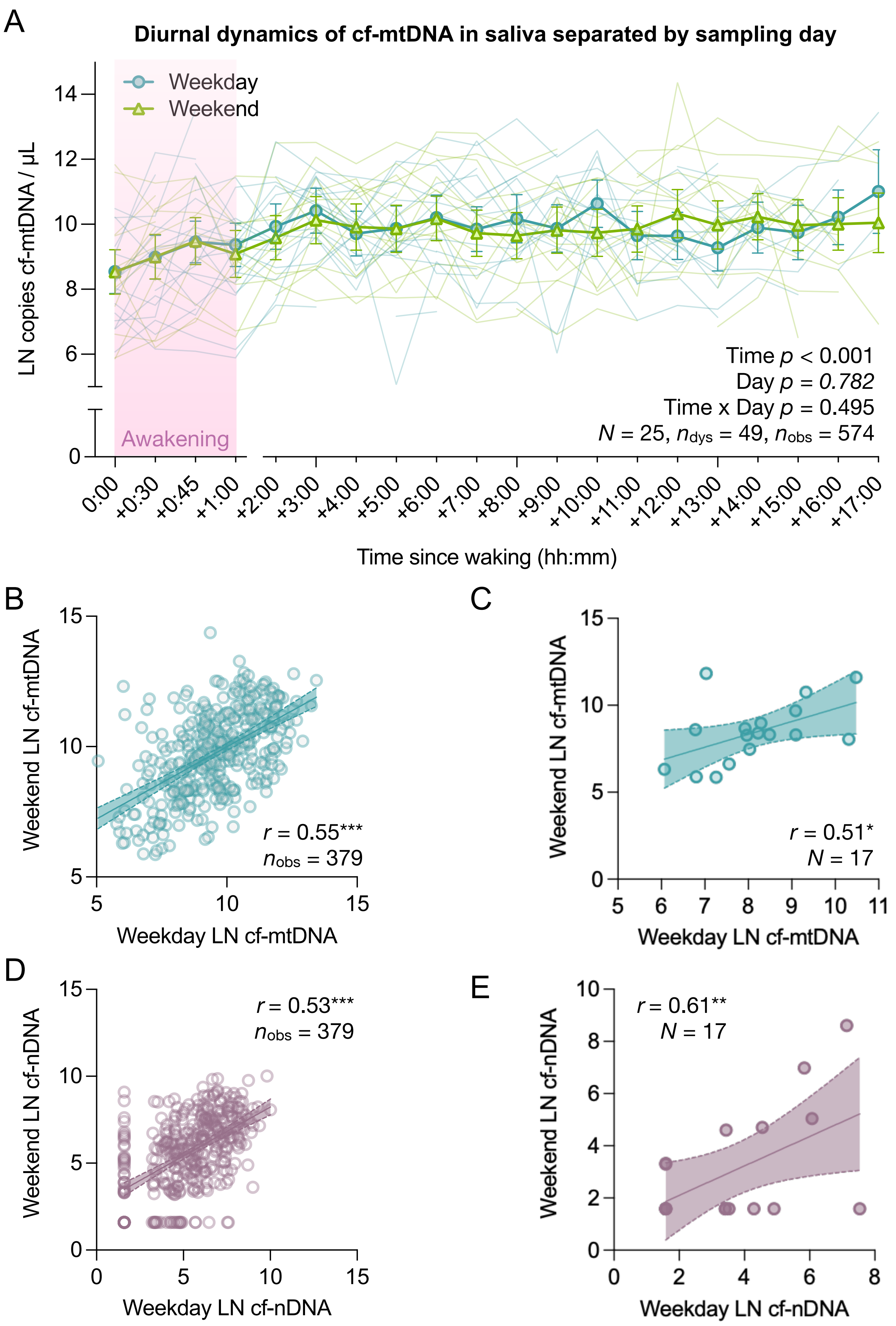


**Supplemental Figure S3. Saliva cell-free DNA trajectories on weekdays and weekend days. (A)** Trajectories of saliva cell-free mitochondrial DNA (cf-mtDNA) measured on weekdays (dark green) and weekends (light green). Values are presented as natural log-transformed concentrations (copies / µl saliva); thin lines represent individual trajectories, dots and error bars indicate linear mixed-effects model-derived marginal means and 95% confidence intervals. Considering the sampling day as main and interaction effect did not improve the model quality (see also Supplementary Table S3). **(B-E)** Scatter plots illustrating the between-day associations of cell-free DNA concentrations in saliva sampled on weekend days and weekdays, with cf-mtDNA concentrations from corresponding time points **(B)** during the entire day and **(C)** upon awakening, and cell-free nuclear DNA (cf-nDNA) concentrations from corresponding time points **(D)** during the entire day and **(E)** upon awakening. Values are presented as (A) marginal means from mixed-effects models with 95% confidence intervals (bold opaque lines and symbols) and individual trajectories (thin transparent lines). Linear trend lines and confidence intervals in (B-E) serve illustrative purposes. Effect sizes and *p*-values from (A) linear mixed-effects models and (B-E) Spearman rank correlations (two-tailed), $^{*}$ $p < 0.05$, $^{**}$ $p < 0.01$, $^{***}$ $p < 0.001$.

Figure S4

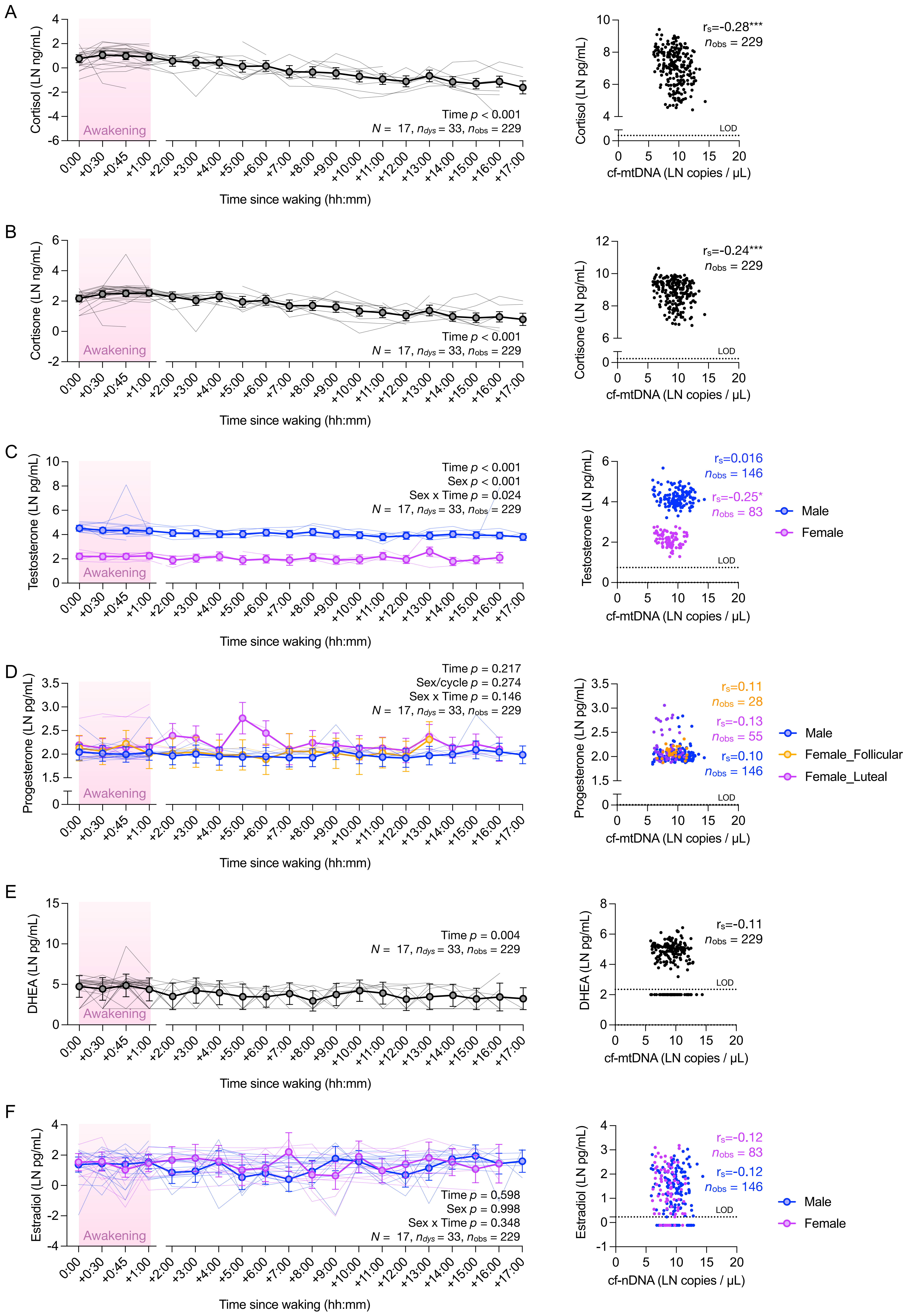
A
Cortisol (LN ng/mL)
Awakening
Time p < 0.001
N = 17, n_dys = 33, n_obs = 229
Time since waking (hh:mm)
0:00 +0:30 +0:45 +1:00 +2:00 +3:00 +4:00 +5:00 +6:00 +7:00 +8:00 +9:00 +10:00 +11:00 +12:00 +13:00 +14:00 +15:00 +16:00 +17:00
Cortisol (LN pg/mL)
rs=-0.28***
n_obs = 229
LOD
cf-mtDNA (LN copies / µL)
B
Cortisone (LN ng/mL)
Awakening
Time p < 0.001
N = 17, n_dys = 33, n_obs = 229
Time since waking (hh:mm)
Cortisone (LN pg/mL)
rs=-0.24***
n_obs = 229
LOD
cf-mtDNA (LN copies / µL)
C
Testosterone (LN pg/mL)
Time p < 0.001
Sex p < 0.001
Sex x Time p = 0.024
N = 17, n_dys = 33, n_obs = 229
Awakening
Time since waking (hh:mm)
Testosterone (LN pg/mL)
rs=0.016
n_obs = 146
rs=-0.25*
n_obs = 83
LOD
Male
Female
cf-mtDNA (LN copies / µL)
D
Progesterone (LN pg/mL)
Time p = 0.217
Sex/cycle p = 0.274
Sex x Time p = 0.146
N = 17, n_dys = 33, n_obs = 229
Awakening
Time since waking (hh:mm)
Progesterone (LN pg/mL)
rs=0.11
n_obs = 28
rs=-0.13
n_obs = 55
rs=0.10
n_obs = 146
LOD
Male
Female_Follicular
Female_Luteal
cf-mtDNA (LN copies / µL)
E
DHEA (LN pg/mL)
Time p = 0.004
N = 17, n_dys = 33, n_obs = 229
Awakening
Time since waking (hh:mm)
DHEA (LN pg/mL)
rs=-0.11
n_obs = 229
LOD
cf-mtDNA (LN copies / µL)
F
Estradiol (LN pg/mL)
Time p = 0.598
Sex p = 0.998
Sex x Time p = 0.348
N = 17, n_dys = 33, n_obs = 229
Awakening
Time since waking (hh:mm)
Estradiol (LN pg/mL)
rs=-0.12
n_obs = 83
rs=-0.12
n_obs = 146
LOD
Male
Female
cf-nDNA (LN copies / µL)

**Supplemental Figure S4. Diurnal trajectories of additional hormones in saliva.** Trajectories of **(A)** cortisol, **(B)** cortisone, **(C)** testosterone, **(D)** progesterone, **(E)** DHEA, and **(F)** estradiol over the diurnal period. Scatter plots to the right of each trajectory illustrate the relation between each hormone and cf-mtDNA. Horizontal dotted lines on scatter plots represent limits of detection (LOD). Values are presented as natural log-transformed concentrations. Thin lines represent individual trajectories. In (A-D), dots and error bars indicate linear mixed-effects model-derived marginal means and 95% confidence intervals, while in (E,F) averaged trajectories are raw data means with standard deviations since the mixed-effects models for DHEA and estradiol did not map the data with sufficient accuracy. Trajectories for (C) testosterone and (F) estradiol are separated by sex, while trajectories for (D) progesterone are separated by sex and menstrual cycle phase. Corticosterone trajectories are not shown due to 90% missing/not-detectable values. Effect sizes and *p*-values from (trajectories) linear mixed-effects models and (scatter plots) Spearman rank correlations (two-tailed), $^{*}$ $p < 0.05$, $^{**}$ $p < 0.01$, $^{***}$ $p < 0.001$.

Figure S5

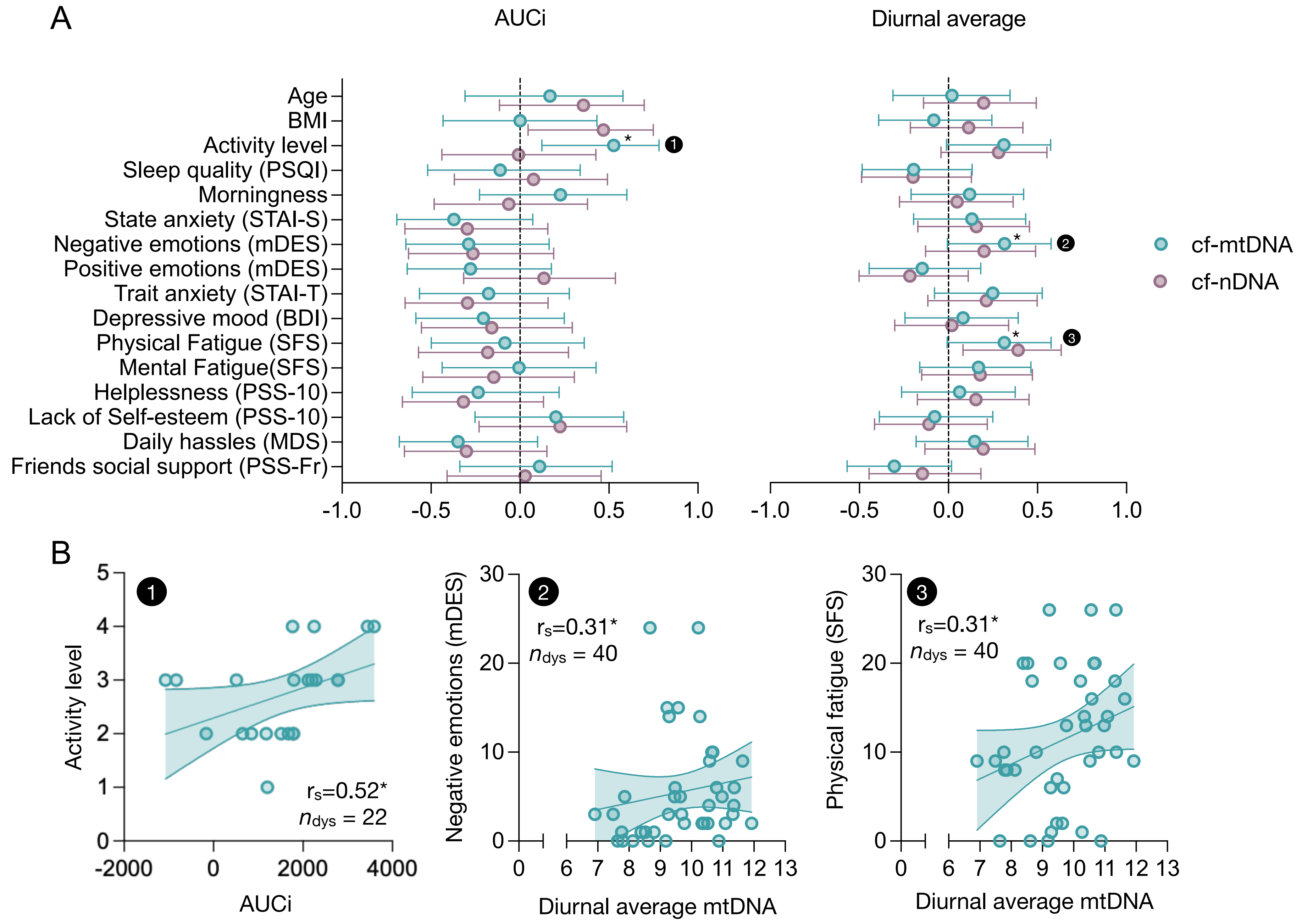


**Supplemental Figure S5. Correlations of saliva cell-free DNA measurements with psychobiological parameters. (A)** Forest plots summarizing the results of Spearman rank correlations between participants' cell-free mitochondrial DNA (cf-mtDNA) and cell-free nuclear DNA (cf-nDNA) diurnal metrics (area under the curve from baseline, AUCi; and diurnal average) and psychobiological characteristics. Dots and error bars indicate effect sizes and 95% confidence intervals of correlations. Encircled numbers indicate significant correlations illustrated as **(B)** scatter plots; linear trend lines and confidence intervals serve illustrative purposes. Effect sizes and two-tailed *p*-values from Spearman rank correlations, $^{*}$ $p < 0.05$, $^{**}$ $p < 0.01$, $^{***}$ $p < 0.001$.
